# Relationships among High School Student-Athletes’ Mental Health, Stressors, and Social Support during the COVID-19 Pandemic in Japan

**DOI:** 10.1101/2021.11.25.21266885

**Authors:** Kaori Yamaguchi, Eriko Katagami, Ryoji Shinohara, Taishi Tsuji, Zentaro Yamagata, Hironobu Tsuchiya

## Abstract

**Objectives:** The impact of the spread of COVID-19 on the mental health and its mitigating factors of high school athletes is not fully understood. The aims of this study were 1) to describe the psychological distress and stressors experienced by high school athletes during the COVID-19 pandemic and to elucidate the relationships between them and 2) to determine the relationship between psychological distress and social support.

**Methods:** Participants of this cross-sectional study were recruited from public high schools in East Japan. We conducted either an online or paper-based questionnaire survey from July 12 to 31, 2020, and used data collected from 3017 high school student athletes (valid response rate: 88.7%) for the analyses. We evaluated psychological distress (K6 ≥10), stressors to athletes during the COVID-19 pandemic (SAC-19), and perceived social support from others. Logistic regression analysis was used to calculate odds ratios (OR) and 95% confidence intervals (CI) for developing psychological distress.

**Results:** Among the participants, 764 (25.3%) experienced psychological distress. Among the five factors extracted from the SAC-19, self-restraint requests (OR = 1.03, 95% CI: 1.01– 1.04), pressure from the surrounding environment (OR = 1.15, 95% CI: 1.12–1.18), and difficulties in maintaining athletic activities (OR = 1.16, 95% CI: 1.12–1.21) increased the risk of psychological distress. On the other hand, participants who were satisfied with the support from family members (OR = 0.77, 95% CI: 0.67–0.90), teammates (the same grade) (OR = 0.81, 95% CI: 0.67–0.98), and coaches and instructors (OR = 0.77, 95% CI: 0.65–0.91) showed lower psychological distress.

**Conclusions:** During the COVID-19 pandemic, high school athletes experienced more psychological distress than usual. Stressors such as self-restraint requests, pressure from the surrounding environment, and difficulties in maintaining athletic activities increased the risk. On the other hand, social support from family members, teammates (the same grade), and coaches and instructors can help alleviate these stressors.

## 1. Introduction

On February 27, 2020, due to the spread of COVID-19, the Japanese government mandated all elementary, junior high, and high schools in the country to close temporarily. Later, on April 16, a nationwide state of emergency was declared due to the rising COVID-19 cases. Children were thus forced to remain at home and suspend their athletic activities for approximately 3 months. Additionally, the various competitions they were eagerly awaiting were cancelled. Therefore, these pandemic-related changes had an impact on their physical and mental health.

The National Center for Child Health and Development^1^ report that during the pandemic, 42% of Japanese high school students experienced recent difficulty in concentrating, 32% had not exercised at least once in the past week, and 26% found that their waking and sleeping hours had shifted by more than 2 hours. These indicate that they were experiencing stress reactions and disruptions in the rhythm of their lives. In addition, Aoki^2^ stated that high school athletes identify club activities as the core and purpose of their lives and are thus crucial determinants of their adjustment and satisfaction in school life. Furthermore, Japanese university students who continue to participate in sports clubs and circles tended to have higher stress-coping abilities and lower psychological distress^3^. Moreover, connecting with others when feeling stressed is considered important to mental health; social support is believed to act as a buffer to mitigate the negative effects of high stress^4^.

Since the outbreak of COVID-19, small-scale school-based surveys of the attitudes of high school athletes have been conducted^5^; however, there have been no large-scale ones assessing for stressors, psychological distress, and stress reactions among high school athletes. Furthermore, research is required to examine what kind of support high school athletes are satisfied with and whether this support alleviate psychological distress and stress responses in situations such as the pandemic.

The purpose of this study is twofold: to understand the psychological distress and stressors experienced by high school athletes during the COVID-19 pandemic and explain the relationship between them; and, to determine the relationship between psychological distress and the level of satisfaction with social support.

Although COVID-19 vaccinations are now available, and certain results have been confirmed regarding the infection, some experts believe that infection prevention practices may continue for several years. Thus, in today’s society, it is important to understand children’s actual state of stress and create strategies to address it in the event of another pandemic, as well as various health crises and large-scale disasters.

## 2. Methods

### 2.1. Study design and participants

This cross-sectional study recruited participants from public high schools in East Japan. High school students enrolled in the extracurricular activity club who provided consented complete the questionnaires (n = 3400; male = 1798; female = 1293). We explained the aims and methods of the study and obtained permission to conduct the survey from the representatives of each school. Additionally, participants were given a digital letter containing a brief description of the study and reassurance of their unrestricted freedom to withdraw participation. The survey was completed either online or using a paper-based questionnaire from July 12 to 31, 2020. Questionnaires with invalid responses, and data from non-athletes were excluded. Thus, data from a final total of 3017 high school student athletes (ages: 15–18 years) were included in the analyses for this study (valid response rate: 88.7%). The present study was approved by the ethics committee of the university with which the last author of this study is affiliated (Approval No. 20-10).

### 2.2. Measures

#### (1) Demographic information

Data on sex, school grade, type of sport played (racket sports, ball games, budo and martial arts, individual sports [e.g., track and field], or dance), and competitive level (regional, prefecture [entry], or prefecture [winners] or higher) were collected.

#### (2) Psychological distress

K6, a scale that measures the degree of mental health problems, was originally developed in English by Kessler et al.^6^, and translated into Japanese by Furukawa et al.^7^ K6 is often used as a screening test for distress experienced in various contexts. The participants rated their perceived stress level within the last 3 months on a 5-point Likert scale ranging from 0 (not all) to 4 (very often). The total possible score ranges within 0–24 points; the higher the score, the worse the individual’s mental health. According to the literature^8,9^, a total score of 10+ on the K6 indicates a risk of mood and anxiety disorders. Therefore, the primary outcome of the present study was set based on this cutoff point.

#### (3) Stress Response Scale for Athletes

We used the Stress Response Scale for Athletes developed by Kemuriyama^10^ as a supplementary outcome of the current study. This scale has five dimensions consisting of 15 items: physical fatigue, apathy, anger, interpersonal distrust, and depression. Participants were asked to recall the frequency of experiencing a stress response during the past 3 months and rated this on a 5-point Likert scale ranging from 1 (not at all) to 5 (very often). A higher score indicates a higher level of stress response.

#### (4) Stressors for athletes during the COVID-19 pandemic

Since no standardized tools are available to assess the pandemic related stressors experienced by student athletes, the stressors for athletes during the COVID-19 pandemic (SAC-19) scale was developed for this study based on previous reports on the stressors or issues faced by athletes or those participating in sports during the COVID-19 pandemic (e.g., National Collegiate Athletic Association Report [NCAA]^11^). The scale consists of 16 items, including stressors related to school (e.g., lack of opportunities to meet classmates), the home (e.g., financial issues within the family), and sporting activities (e.g., cancellation or suspension of competitions). Participants rated the items on a 5-point Likert scale ranging from 1 (not at all) to 5 (very often); the total score ranges from 16 to 80.

#### (5) Social support for athletes during the COVID-19 pandemic

Perceived social support consists of six items measuring satisfaction with the support received from 1) family, 2) classmates and friends, 3) teachers, 4) same grade teammates, 5) teammates from other grades, and 6) coaches and instructors during the COVID-19 pandemic. Participants rated these items on a 5-point Likert scale ranging from 1 (not at all satisfied) to 5 (very satisfied) based on the past 3 months. We dichotomized scores of 4 and 5 as “satisfied” (1) and scores of 1 to 3 as “not satisfied” (0).

### 2.3. Statistical analysis

We performed descriptive statistical analysis of the participants according to sex, competitive level, and school grades. The unpaired t-test and one-way analysis of variance were used to compare continuous variables, whereas the chi-square test was used for categorical variables. We conducted exploratory factor analysis using the maximum likelihood method and oblimin rotation to examine the factorability of the SAC-19. We determined the number of factors using a scree plot and assessed the internal consistency by calculating the Cronbach’s alpha coefficient. We performed multivariable Poisson regression analysis to examine the association of demographic characteristics, stressors for athletes, and social support with psychological distress (K6 ≥ 10). When the incidence of an outcome of interest is common in the study population (>10%), the adjusted odds ratio derived from the logistic regression can no longer approximate the risk ratio^12^. The prevalence of psychological distress in the present study was 25.3%; therefore, we applied Poisson regression and calculated the prevalence ratio (PR) and its 95% confidence interval (CI). Three models were included the following: Model 1, demographic characteristics (sex, school grade, type of sports, and competitive level); Model 2, Model 1 + each factor of the SAC-19; and Model 3, Model 2 + social support. As a sensitivity analysis, we performed multiple regression analysis with the total score of the Stress Response Scale for Athletes as an outcome and constructing the same three models. Stata/MP 16.1 (StataCorp, College Station, TX, USA) was used to calculate descriptive statistics and performing multivariable regression analyses, with *P* <.05 indicating statistical significance. SAS 9.4 software (SAS Institute Inc., Cary, NC, USA) was used for the exploratory factor analysis.

## 3. Results

Table 1 presents the descriptive characteristics of the participants. Among the 3017 participants, 764 (25.3%) experienced psychological distress. Approximately 60% to 70% of the participants were satisfied with each type of social support. Exploratory factor analysis of SAC-19 indicated five factors composed of the 16 variables (Table 2). They were labeled “restriction on sport activities (F1),” “self-restraint request (F2),” “fear of exposure to COVID-19 (F3),” “pressure from the surrounding environment (F4),” and “difficulties in maintaining athletic activities (F5).” The Cronbach’s alpha coefficients were 0.92, 0.83, 0.87, 0.55, and 0.68 for F1, F2, F3, F4, and F5, respectively. Furthermore, the coefficient for the overall scale was 0.91, thus indicating the adequate internal consistency of the scale. Female participants scored higher on the Stress Response Scale for Athletes, as well as on F2 and F3 components of the SAC-19 (Supplementary Table 1). Those with higher competitive levels had higher scores in F1, F2, F4, and F5 (Supplementary Table 2). Psychological distress, stress response, F1, F2, F4, and F5 were more prevalent among students from higher school grades (Supplementary Table 3).

**Table 1.** Descriptive characteristics of the participants (n = 3017)

| Variables |  |  |
| --- | --- | --- |
| **Sex (n, %)** |  |  |
| Male | 1726 | 57.2% |
| Female | 1291 | 42.8% |
| **School grade (n, %)** |  |  |
| 1st grade | 1274 | 42.2% |
| 2nd grade | 1221 | 40.5% |
| 3rd grade | 522 | 17.3% |
| **Type of sports (n, %)** |  |  |
| Racket sports | 882 | 29.2% |
| Ball games | 1310 | 43.4% |
| Budo and martial arts | 216 | 7.2% |
| Individual sports (e.g., track and field) | 569 | 18.9% |
| Dance | 40 | 1.3% |
| **Competitive level (n, %)** |  |  |
| Regional | 1253 | 41.5% |
| Prefecture (entry) | 1120 | 37.1% |
| Prefecture (winners) or higher | 644 | 21.3% |
| **Psychological distress (n, %)** |  |  |
| Kessler-6 score $\geq 10$ | 764 | 25.3% |
| **Stress response (mean, SD)** |  |  |
| Stress Response Scale for Athletes | 65.5 | 21.0 |
| **SAC-19 (mean, SD)** |  |  |
| Restriction on sport activities | 16.4 | 7.3 |
| Self-restraint request | 11.7 | 4.5 |
| Fear of exposure to COVID-19 | 7.3 | 3.3 |
| Pressure from the surrounding environment | 6.3 | 2.5 |
| Difficulties in maintaining athletic activities | 5.1 | 2.2 |
| **Social support, satisfied (n, %)** |  |  |
| Family | 2143 | 71.0% |
| Classmates and friends | 2181 | 72.3% |
| Teachers | 1767 | 58.6% |
| Teammates (the same grade) | 2157 | 71.5% |
| Teammates (other grades) | 2098 | 69.5% |
| Coaches and instructors | 1914 | 63.4% |

SAC-19: Stressors for Athletes during the COVID-19 pandemic.

**Table 2.**
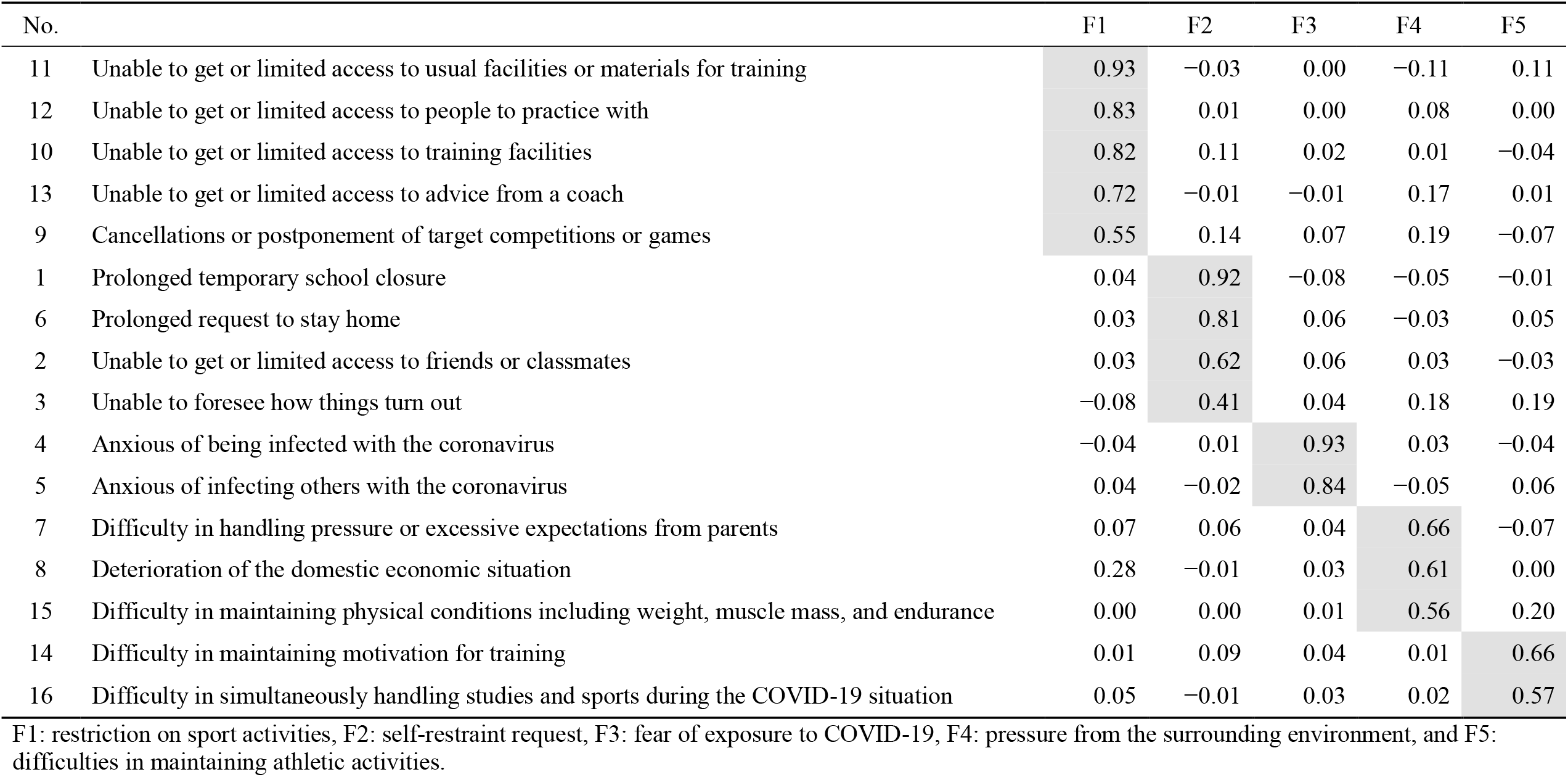
Exploratory factor analysis for the Stressors for Athletes during the COVID-19 pandemic

Table 3 shows the results of the multivariable Poisson regression analysis elucidating the association of demographic characteristics, stressors for athletes, and social support with psychological distress. In Model 1, the prevalence of psychological distress was higher in 2nd and 3rd grade students than 1st grade students, and lower for participants at the prefecture competition (entry) level than those at the regional competition level. In Model 2, F4 and F5 were positively associated with psychological distress, whereas F1 was negatively associated. Model 3, which introduced social support, showed that participants who were satisfied with the support from family members, teammates (the same grade), and coaches and instructors had a lower frequency of psychological distress than those who did not. Similar results were obtained from the sensitivity analysis using the Stress Response Scale for Athletes as an outcome (Supplementary Table 4).

**Table 3.**
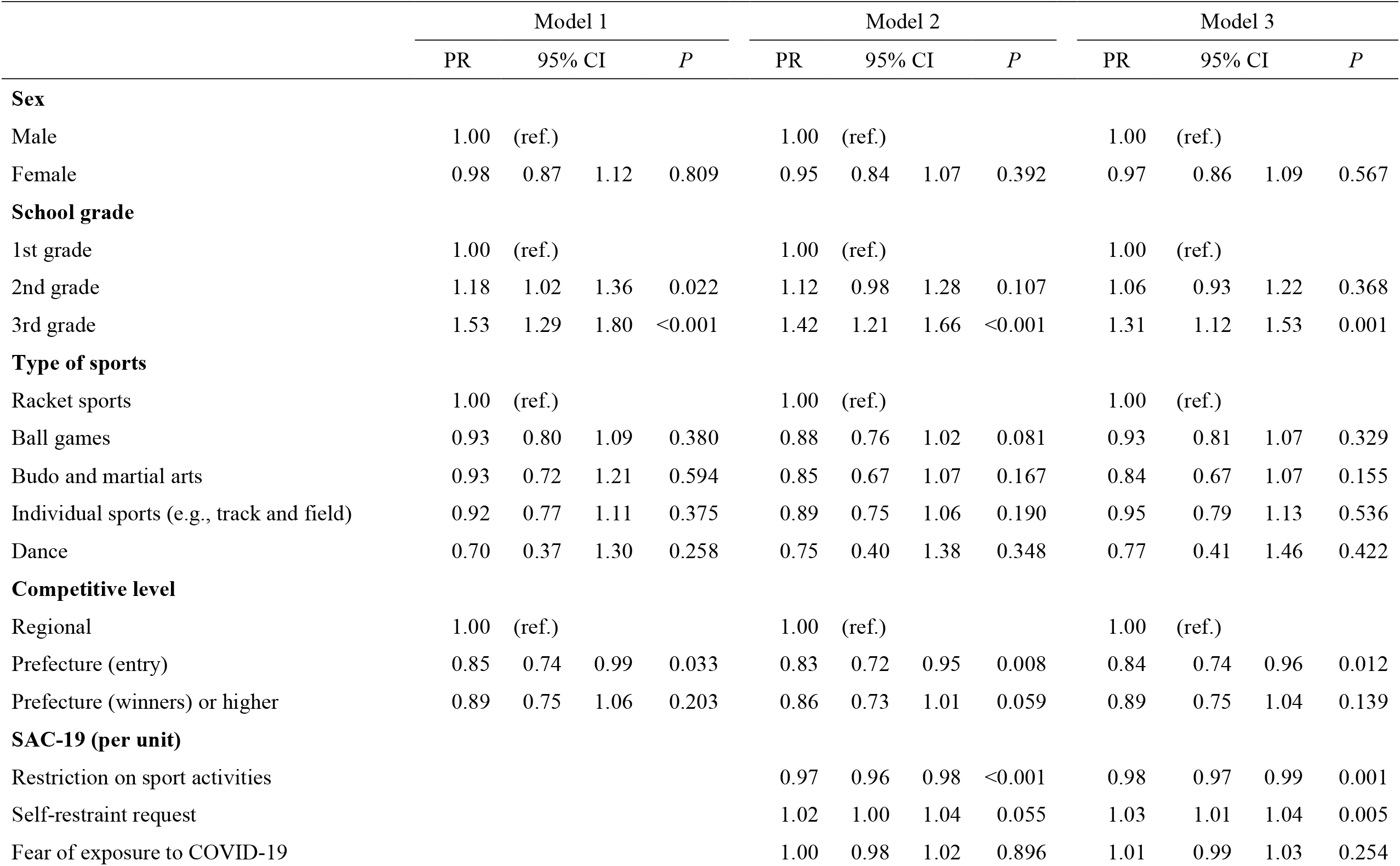
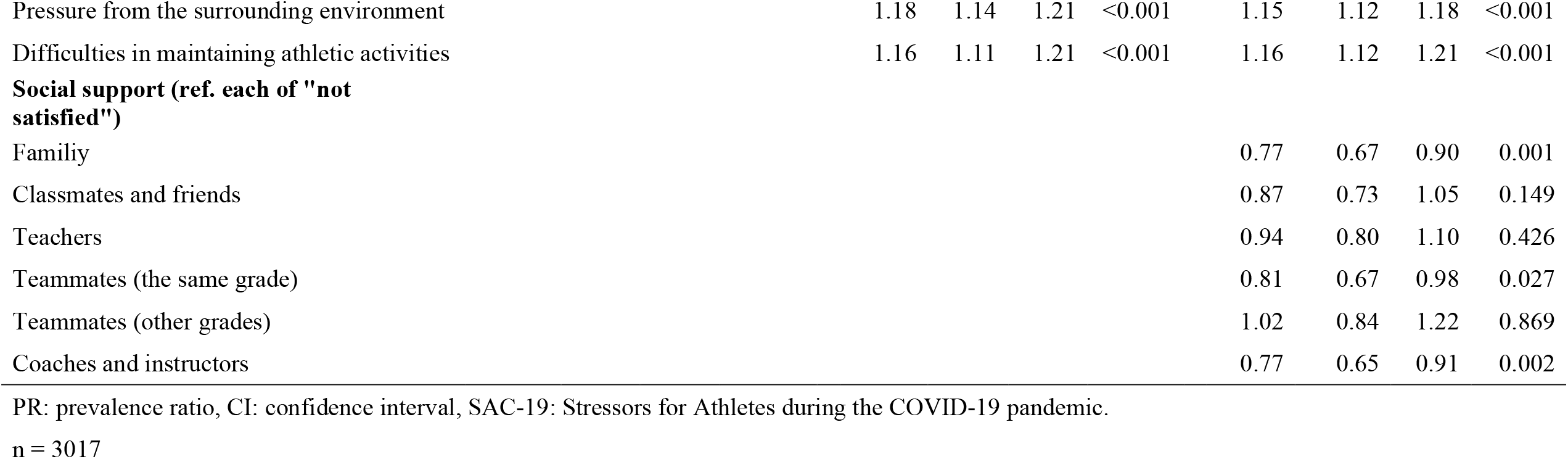
Associations between psychological distress (Kessler-6 with a cut-off score of 9/10) and demographic characteristics, stressors for athletes, and social support

## 4. Discussion

In this study, we assessed the relationship between psychological distress and stressor status among high school athletes during the COVID-19 pandemic. We then examined whether the psychological distress status differed depending on the level of satisfaction with support from the surrounding environment. The results showed that approximately 25% of the participants experienced strong psychological distress. The risk was increased by the following stressors: self-restraint request, pressure from the surrounding environment, and difficulties in maintaining athletic activities. Meanwhile, those who were satisfied with support received from family members, teammates (the same grade), and coaches and instructors showed lower psychological distress.

### 4.1. Psychological distress faced by high school athlete during the COVID-19 pandemic

Our findings point to the serious deterioration of mental health among high school student athletes due to the pandemic. In a survey conducted before the pandemic with participants in a similar age group (12–19 years)^13^, 7.8% of the participants scored 10+ points on the K6 score, while 25.3% of the sample scored it in the current study. Thus, the need for providing care and support to high school student athletes is evident.

The pandemic has created numerous challenges for elite athletes^14^ and for the world of sport in general^15,16^. According to the Athlete 365 survey findings^17^, athletes reported facing challenges in keeping up with regular effective training and in staying motivated. Managing their mental health and sporting careers were reported as challenges. Financial concerns associated with continuing their sporting careers were also reported as a critical issue, especially for elite athletes. Not only in elite sports, but also athletes across a wide range of categories faced such difficulties due to the pandemic-related restrictions. Similarly, the temporary closure of high schools and restrictions on training duration for sporting activities led to stress and difficulty for high school student athletes.

### 4.2. Stressors to athletes during the COVID-19 pandemic

The SAC-19, which was mainly developed for this study, was divided into factors as expected, and the Cronbach’s alpha was sufficiently high, indicating acceptable internal consistency. Regarding sex differences, female athletes had higher stress scores with self-restraint request (F2) and fear of exposure to COVID-19 (F3) than male athletes. According to the Ministry of Health, Labour and Welfare’s patient survey^18^, the number of depressed patients was higher among females in all age groups, ranging from teens to 80s. The NCAA survey^11^ also reported that female athletes were more concerned than male athletes of approximately 9 out of 10 mental health concerns, and the results of the present study support this. The higher the school grades and competition levels, the more significant the stressors for the restriction of athletic activities (F1), self-restrained life (F2), pressure from the surrounding environment (F4), and difficulties in maintaining athletic activities (F5). Normally, the higher the school grades and competition levels, the more opportunities to participate and play an active role in competitions. This situation could lead to higher self-expectations. In addition, for 3rd grade students, there may be a relation to the influence of competition results on their career paths.

### 4.3. Relationship of psychological distress with stressors and demographic characteristics

Self-restraint request (F2) was associated with high level of psychological distress. With the cessation of sports activities, the participants had more time to spare but were unable to go out or meet with friends. Living at home for a long time must have been boring and stressful. In the same way, the pressure from the surrounding environment (F4) and difficulties in maintaining athletic activities (F5) were positively associated with psychological distress, which is in line with our hypothesis. The NCAA survey^11^ also reported that family financial situations negatively affected the mental health of athletes.

Meanwhile, restriction on sport activities (F1) was inversely related to psychological distress. Although elucidating the reasons is difficult, high school-age students have various interests other than academics and sports. The suspension of activities possibly created a sense of freedom from sports for a while. However, no significant association was found between psychological distress and fear of exposure to COVID-19 (F3), which may be because COVID-19 is not easily transmitted to young people, and, even when infected, it is not likely going to be serious; additionally, few people of the same generation in their surroundings were infected.

We observed that the prevalence of psychological distress was higher among those in higher school grades. This can be attributed to the same reasons mentioned above. No significant differences were found between the types of sports. Before the survey, we speculated that contact sports, which are risk factors for COVID-19, might have a large impact. However, all sports regardless of the type would have been affected in a similar manner. For competition level, we hypothesized that the higher the competition level, the greater the impact on psychological distress, however, no difference was found. Motivation to engage in sports is highly individualized, and probably many factors other than the level of competition exist; this requires further research.

### 4.4. Relationship between psychological distress and social support

Previous studies of college athletes during the COVID-19 pandemic have reported that social support from teammates was positively associated with mental health^19,20^. We also found that social support from teammates, especially from the same grade, is expected to be effective, whereas support from the surrounding adults, such as family members and coaches and instructors, is also important. During the self-restraint period, the students spent a lot of time with their family members, indicating that they felt pressure from their families in some areas but that they trusted them. In addition, teammates seemed to provide a space for them to share their anxieties and worries. Many of the coaches and instructors supported the students during the self-restraint period by meeting online and sending text messages^21^. This kind of support from people with close ties to the respondents seemed to be effective.

### 4.5. Strengths and limitations

The strength of the current study was the use of data from a large-scale survey of high school athletes during the COVID-19 pandemic. This is not only the first survey in Japan, but also provide valuable data that are applicable worldwide. However, this study has several limitations. First, generalizability is not sufficiently high. The survey was conducted in a relatively limited area in Japan, and since the infection status and behavioral restrictions differed depending on the period, the results possibly differed depending on the survey conditions. Furthermore, restrictions on activities may differ depending on the school, and the extent to which the activities were restricted is unknown. Second, the cross-sectional study design does not allow for a discussion of causal relationships. Third, the criterion-related validity of the SAC-19 against the gold standard items were not confirmed in advance because emphasis was placed on conducting a rapid investigation. However, the internal consistency was acceptable, and the results that were in line with our hypotheses were confirmed in the current study; therefore, this might not be a major limitation.

## 5. Conclusions

During the COVID-19 pandemic, approximately 25% of high school athletes experienced stronger psychological distress than usual. Stressors such as self-restraint request, pressure from the surrounding environment, and difficulties in maintaining athletic activities increased the risk. Social support from family members, teammates (the same grade), and coaches and instructors alleviated these stressors.

A study conducted by Marani et al.^22^ shows that pandemics such as COVID-19 are not rare and may occur more frequently in the future. The probability of a pandemic having the same impact as COVID-19 is approximately 2% per year. In addition to pandemics, large-scale disasters are not uncommon in Japan. This study provides an important basis for understanding how young athletes are stressed and their individual and social support needs for coping in such emergencies.

## Data Availability

The data that support the findings of this study are available from the corresponding author, KY, upon reasonable request.

